# Implementation of Smart Triage combined with a quality improvement program for children presenting to facilities in Kenya and Uganda: An interrupted time series analysis

**DOI:** 10.1101/2024.02.09.24302601

**Authors:** J Mark Ansermino, Yashodani Pillay, Abner Tagoola, Cherri Zhang, Dustin Dunsmuir, Stephen Kamau, Joyce Kigo, Collins Agaba, Ivan Aine Aye, Bella Hwang, Stefanie K Novakowski, Charly Huxford, Matthew O. Wiens, David Kimutai, Mary Ouma, Ismail Ahmed, Paul Mwaniki, Florence Oyella, Emmanuel Tenywa, Harriet Nambuya, Bernard Opar Toliva, Nathan Kenya-Mugisha, Niranjan Kissoon, Samuel Akech, the Pediatric Sepsis CoLab

**Affiliations:** Department of Anesthesiology, Pharmacology & Therapeutics, University of British Columbia, 217-2176 Health Sciences Mall, Vancouver, BC V6T 1Z3; Institute for Global Health, BC Children’s Hospital and BC Women’s Hospital + Health Centre, 305-4088 Cambie Street, Vancouver, BC, V5Z 2X8; Department of Pediatrics, Jinja Regional Referral Hospital, Jinja, Uganda; KEMRI-Wellcome Trust Research Programme, P.O. Box 43640-00100, Nairobi, Kenya; World Alliance for Lung and Intensive Care Medicine in Uganda, Plot 5-7, Coral Crescent, Kololo, P.O. Box 9924, Kampala, Uganda; Department of Paediatrics, Mbagathi County Hospital, Nairobi County, Kenya; Department of Pediatrics, Gulu Regional Referral Hospital, Gulu, Uganda; Department of Pediatrics, University of British Columbia, Rm B2W, 4480 Oak Street, Vancouver, BC, V6H 3V4

## Abstract

Sepsis occurs predominantly in low-middle-income countries. Sub-optimal triage contributes to poor early case recognition and outcomes from sepsis. We evaluated the impact of Smart Triage using improved time to intravenous antimicrobial administration in a multisite interventional study.

Smart Triage was implemented (with control sites) in Kenya (February 2021-December 2022) and Uganda (April 2020-April 2022). Children presenting to the outpatient departments with an acute illness were enrolled. A controlled interrupted time series was used to assess the effect on time from arrival at the facility to intravenous antimicrobial administration. Secondary analyses included antimicrobial use, admission rates and mortality (NCT04304235).

During the baseline period, the time to antimicrobials decreased significantly in Kenya (132 and 58 minutes) at control and intervention sites, but less in Uganda (3 minutes) at the intervention site. Then, during the implementation period in Kenya, the time to IVA at the intervention site decreased by 98 min (57%, 95% CI 81-114) but increased by 49 min (21%, 95% CI: 23-76) at the control site. In Uganda, the time to IVA initially decreased but was not sustained, and there was no significant difference between intervention and control sites. At the intervention sites, there was a significant reduction in IVA utilization of 47% (Kenya) and 33% (Uganda), a reduction in admission rates of 47% (Kenya) and 33% (Uganda) and a 25% (Kenya) and 75% (Uganda) reduction in mortality rates compared to the baseline period.

We showed significant improvements in time to intravenous antibiotics in Kenya but not Uganda, likely due to COVID-19, a short study period and resource constraints. The reduced antimicrobial use and admission and mortality rates are remarkable and welcome benefits but should be interpreted cautiously as these were secondary outcomes. This study underlines the difficulty of implementing technologies and sustaining quality improvement in health systems.

**Author Summary:** Implementing the Smart Triage platform and quality improvement program for children in Kenya and Uganda resulted in inconsistent improvements in time to intravenous antimicrobial administration. The time to IVA decreased significantly in Kenya during baseline and reduced further during the intervention while increasing at the control site. In Uganda the time to treatment initially decreased but was not sustained. The treatment times were significantly influenced by the improvements during baseline data collection and multiple external health system factors such as drug shortages, the COVID -19 pandemic, staff shortages and strikes. The dramatic reduction in treatment, admission, and mortality rates should be further investigated.

## Background

Among critically ill children, sepsis is the leading cause of preventable deaths globally. Countries in Sub-Saharan Africa report disproportionately high case fatality rates (1, 2). Many of these deaths are from malaria, pneumonia and diarrheal diseases resulting in sepsis, which may respond to time-sensitive treatment (3). Early treatments, usually within the first hour of presentation, improve survival rates (4, 5). However, delayed recognition and treatment within health facilities remain significant barriers to reducing sepsis-related deaths and complications (6).

Critically ill children can be rapidly identified using triage, which prioritizes patients and the provision of medical care according to illness severity (7). The Emergency Triage Assessment and Treatment (ETAT) system (8, 9), the Pediatric South African Triage Scale (PSATS) (10) and Integrated Management of Childhood Illnesses (IMCI) guidelines (11) were developed based on expert consensus to facilitate triage processes for low-resourced environments. Despite adoption and scaling in Lower-and-Middle-Income Countries (LMICs), sustained implementation into clinical practice at scale has been a persistent challenge, resulting in suboptimal impact (9, 12, 13). Therefore, triage remains under-used in many LMIC settings and patients are still frequently seen on a first-come-first-served basis (14).

Recent efforts to improve triage and treatment decisions include clinical decision support tools implemented in digital platforms using guidelines based on expert opinion (15). Digital tools can also provide automated guidance based on real-world data using an individualized precision public health approach (15) and data-driven feedback that can be used for quality improvement (QI) by health workers (16) and to address local implementation challenges (17). Nevertheless, multifaceted system-wide interventions are still needed to effectively improve care practices in complex, low-resourced settings (3).

We have developed, validated, implemented, and evaluated a digital triage platform called Smart Triage (18–22). The Smart Triage platform comprises data-driven risk assessment (triage), patient and treatment tracking, a real-time dashboard, and automated reports to improve emergency case recognition and time-to-treatment. The Smart Triage algorithm predicts admission, as a data-driven surrogate for illness severity, and is combined with emergency and priority signs as guardrails to generate a triage tool suitable for all acutely ill patients.

QI studies evaluating digital triage tools in low-income countries, under real-world conditions, are scarce (13). We report on the findings of a study investigating the Smart Triage platform implementation at pediatric outpatient departments (OPD) of four tertiary hospitals (two hospitals in Uganda and two hospitals in Kenya). We hypothesized that the Smart Triage platform would reduce the time to treatment. The secondary aims were to evaluate the effect of Smart Triage and QI on patient outcomes, including mortality, admission and readmission.

## Methods

We use an interrupted time series analysis to assess changes in the time from arrival at the facility to administration of an appropriate sepsis bundle of care following implementation of the Smart Triage platform.

### Study sites

This controlled interrupted time series study was conducted in the OPDs of four public tertiary facilities in Kenya and Uganda. Facilities included one control and one intervention site per country. In Kenya, the facilities were Mbagathi County Hospital (intervention site) and Kiambu County Referral Hospital (control site). In Uganda the facilities were Jinja Regional Referral Hospital (intervention site) and Gulu Regional Referral Hospital (control site) (Fig 1).

**Figure 1.**
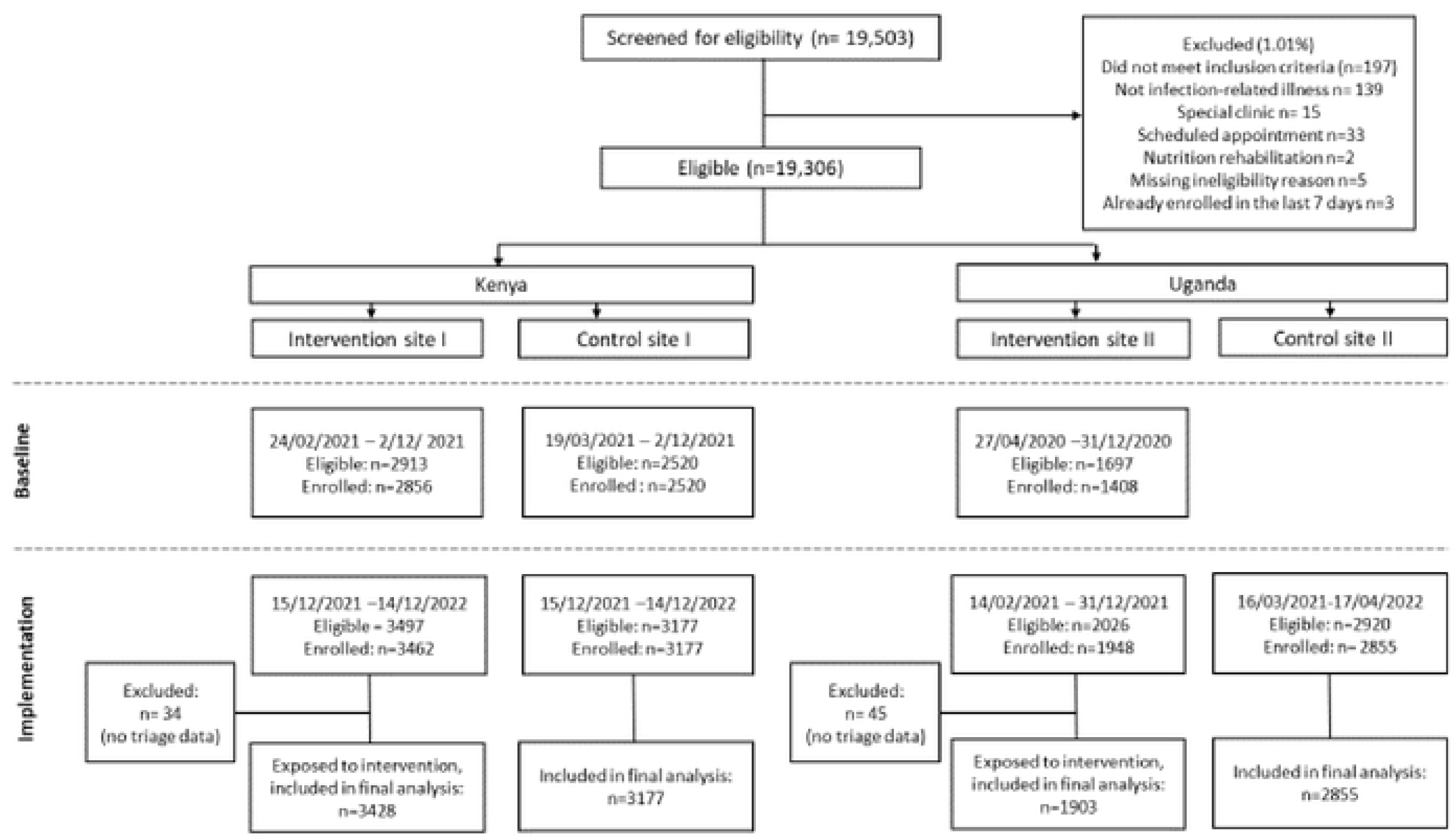
Flow diagram of the Smart Triage multisite interventional study in Uganda and Kenya indicating the progress through study phases at each site. Control site was added later than intervention site in Kenya due to site initiation delays in intervention site and control site in Uganda resulting from the COVID-19 pandemic.

### Contextual and temporal factors

COVID-19-related lockdowns restricting gatherings and travel between districts were enforced from 15 April-5 May 2020 and 7 June-19 July 2021 in Uganda. Outside of these dates, a strict curfew and other infection prevention measures were observed from April 2020 to December 2021. The study was initiated in Kenya after in-country COVID-19-related restrictions had ended. In addition, there were healthcare worker shortages due to strikes (nurses in Kenya and junior doctors in Uganda). Planned staff rotations also significantly impacted compliance with triage and QI. The control site in Uganda was added a year later than the intervention site in Uganda due to delays in study initiation at the control site in Kenya, which was initially planned as the sole control site.

### Participant recruitment

All pediatric arrivals presenting with an acute illness to the OPD were eligible. The age limits were in keeping with each hospital’s practice for pediatric admissions. Informed consent was obtained from parents or legal guardians. Assent was obtained from children over 8 years (Uganda) and 13 years (Kenya). We excluded trauma, burns, elective cases (surgery, change of dressing), immunization visits or clinical review or follow-up appointments.

### Standard Procedures

The implementation design and procedural information have previously been published (20). In brief, a preintervention phase (baseline), interphase (model and technology development) and intervention phase (implementation) were used. The implementation is described in this report. We used a systematic method of sampling. Following consent, predefined candidate predictor variables and outcomes were collected on patients waiting to be seen in the OPD by trained study nurses using Android tablets on a custom-built mobile application (18, 20). Dedicated timekeepers tracked time to treatments. The implementation and control sites continued data collection for the entire study duration. Oxygen saturation (SpO2) and heart rate were measured using the Masimo iSpO2® (Masimo Corporation, California, USA). Respiratory rate was measured using the RRate Application directly on the tablet (23, 24). The Welch Allen SureTemp 692 thermometer was used to measure temperature (Welch Allyn, New York, USA).

### Smart Triage Platform

Using the baseline data at intervention sites, we developed and recalibrated a nine-variable risk prediction model (19) and deployed this within a mobile application for triage. The triage algorithm included independent triggers based on the ETAT triage guidelines. The independent triggers ensure that no danger or priority sign or single significantly abnormal vital sign is missed. The Smart Triage platform comprises data-driven risk assessment (triage), patient and treatment tracking using a Bluetooth Low Energy (BLE) tracking system called Smart Spot, a real-time dashboard, and automated reports. The Smart Spot tags communicated with readers strategically located in the hospital to provide the patient’s location as they move through the facility. Treatment tags in the treatment or emergency room were used to track the time and type of treatment given. All the information collected at triage and via the Smart Spot system was accessible through the clinical dashboard. The dashboard was available to clinicians, laboratory, pharmacy, nutrition team and administrators on laptops, tablets or computer screens. A large screen with a public-facing dashboard was displayed in the waiting areas. The public-facing dashboard displayed coded patient queues, health information posters and videos targeting caregivers. The information was intended to improve understanding of the triage process, Smart Spot system and general health information.

The feasibility and usability of the platform were assessed through questionnaires, direct observation and usability scenarios using a think-aloud methodology (20). Qualitative interviews were used to optimize the platform (21). A cost-effectiveness study was completed to determine utilization costs and implications for scale-up (22).

### Implementation

Hospital triage staff routinely utilized the digital triage platform during the intervention phase. Following triage with Smart Triage algorithm, families were given colour-coded lanyards, which corresponded to the predicted level of risk, with an attached Smart Spot BLE tag. Red lanyards corresponded to emergency cases, yellow to priority and green to non-urgent cases.

The Smart Triage platform was implemented as part of the overall QI program that focused on delivering a sepsis bundle of care. At implementation sites, healthcare workers were trained on the importance of early sepsis recognition and treatment, in using the platform and in performing QI using a train-the-trainer model. Weekly feedback reports were provided using data collected by the Smart Triage platform. The reports included information on the number of children triaged, triage categories, duration of triage, treatment times and metrics of specific interest to the facility. The metrics and report layout were developed collaboratively with facility leadership and implementors to improve data-driven QI processes at the facility. Ongoing training, support supervision, job aides and manuals were also available to health workers.

QI initiatives were aimed at optimal triage and time to antimicrobial administration and were led and customized by each implementation facility. Triage QI initiatives included reducing the time from arrival to triage, improving the triage completion rate, increasing completeness of vital sign measurement, and creating a daily updated list of drug and supply stock-outs to improve communication between departments and caregivers. The causes of delayed time to treatment were identified by using a fishbone diagram. The workflow was optimized at both sites with changes in patient queuing at admission, assessment, laboratory, pharmacy and the emergency room. Specific benches were allocated for emergency cases. An emergency pharmacy cabinet with after-hours access was installed in Uganda to reduce the impact of delays in supplying drugs from the pharmacy. Stock-outs and staff shortages were the most significant barriers identified at both sites (Table S1).

### Follow-up

In-hospital and post-discharge mortality and readmission data were collected during a phone call at seven-days post discharge (or post-visit for those who were not admitted).

### Outcome Measures

The primary outcome was the time to IVA from the time of OPD arrival, among those treated with IVA. The initially planned primary outcome was the time to receive an appropriate sepsis bundle of care (antibiotics, antimalarials, oxygen or fluids if clinically indicated). However, it became evident that IVA was most robustly collected by the timekeepers and most commonly occurred after the other elements of the bundle.

Secondary outcomes included treatment rates, the admission rate, readmission rate (both among those admitted and among those not initially admitted), length of stay among admissions, and overall mortality at seven days. In addition, the time to IVA in each triage category and the proportion of triaged patients was calculated.

### Data Management

All data was captured digitally, either on a tablet or through the study dashboard. At the end of each day, data was uploaded directly to REDCap (25), on a Kenya Medical Research Institute (KEMRI) server in Kenya and on the central study server at the BC Children’s Hospital Research Institute for Uganda.

### Sample size

Based on the feasibility study (22), the increase in the proportion of children receiving a bundle of care within one hour in this study was 21.4%, and the proportion of children receiving IVA was 8.2%. Based on these observations, we estimated that using an alpha of .05, at a power of 80%, we would require 10 600 triages (1290 treatments) in the pre-intervention and post-intervention phases combined, to confirm this effect observed in the previous study.

### Analysis

We used Mann-Whitney and Kruskall-Wallis (for continuous variables) and Fischer’s exact test (for categorical variables) to compare patient characteristics for patients who received IVA and outcomes between control and intervention sites at baseline and following implementation. A p-value of <0.001 was considered statistically significant.

We used a quasi-experimental interrupted time series to assess the immediate effects and effects over time of the intervention but excluding the interphase period. The post-implementation period was set to occur the day following the baseline period. Quantile regression was used to estimate the change in median time to IVA (in minutes) per increase in days since enrollment. The independent variable included each individual IVA administration while a weekly median IVA was derived to visually summarize data. The regression error terms were fitted using an Ordinary Least Square method. Separate regression analysis was done for each of the control and intervention sites. We estimated secular trends for the pre- and post-implementation period and compared the post-implementation period with the counterfactual trend to determine immediate and longer-term effect of the implementation using slope and level changes, respectively. An interaction term with sites was fitted to estimate the difference in median time to IVA between control and intervention sites.

A univariate logistic regression was fitted to obtain the odds ratio of admission, readmission, mortality, and receiving IVA between sites at baseline and following implementation as well as within sites at the intervention sites for the full cohort and separately by triage categories. Confidence intervals were calculated using maximum likelihood.

### Ethical Considerations

Ethics approval was obtained from Makerere University School of Public Health (MUSPH) Institutional Review Board (IRB00011353). The Ugandan National Council for Science and Technology (UNCST) (HS528ES) in Uganda provided approval for study activities to be conducted at Jinja Regional Referral Hospital and Gulu Regional Referral Hospital. The KEMRI Scientific and Ethics Review Unit (SERU) (KEMRI/SERU/CGMR-C/183/3958) provided approval for the study activities to be conducted at Mbagathi County Hospital and Kiambu County Hospital in Kenya. The study was also approved by the University of British Columbia Research Ethics Board in Canada (H19-02398-A006). The trial was registered on Clinical Trials.gov. Identifier: NCT04304235, Registered 11 March 2020.

## Results

There were 6784 children enrolled at baseline from three sites and 5410 at two intervention sites and 6032 at two control sites during the implementation phase (Figure 1). There were a few negative times as patients were identified as emergency cases and treated as soon as they arrived before triage. In such cases, negative times were set to zero, and times greater than 8 hours were censored at 8 hours. Triage was completed on 5331 (98.5%) children enrolled at the intervention sites during the implementation period, which formed our analysis cohort. The median (IQR) time for fluids and/or oxygen to be initiated was 70 (32–137) min compared to IVA at 210 (130–304) min at all sites during both phases (Table S2, S3). In addition, only 85 (13.6%) children received fluids or oxygen after IVA. Time to IVA among children who went on to receive IVA was thus used as the study endpoint for all analyses based on these observations.

In Kenya, the baseline time to IVA decreased by 133 min (44%, 95% CI: 93 to 171) at the control site and by 57 min (37%, 95% CI: 38 to 77) at the intervention site (Figure 2). Paradoxically, there was a 74-min (76%; 95% CI: 54-94) increase in time to IVA following the 2-week interphase period at the Kenya intervention site. During the implementation phase, the time to IVA decreased by 98 min (57%, 81-114) at the Kenya intervention site, while during the same period it increased by 50 min (21%, 95% CI: 23-76) at the control site (Table 1).

**Figure 2.**
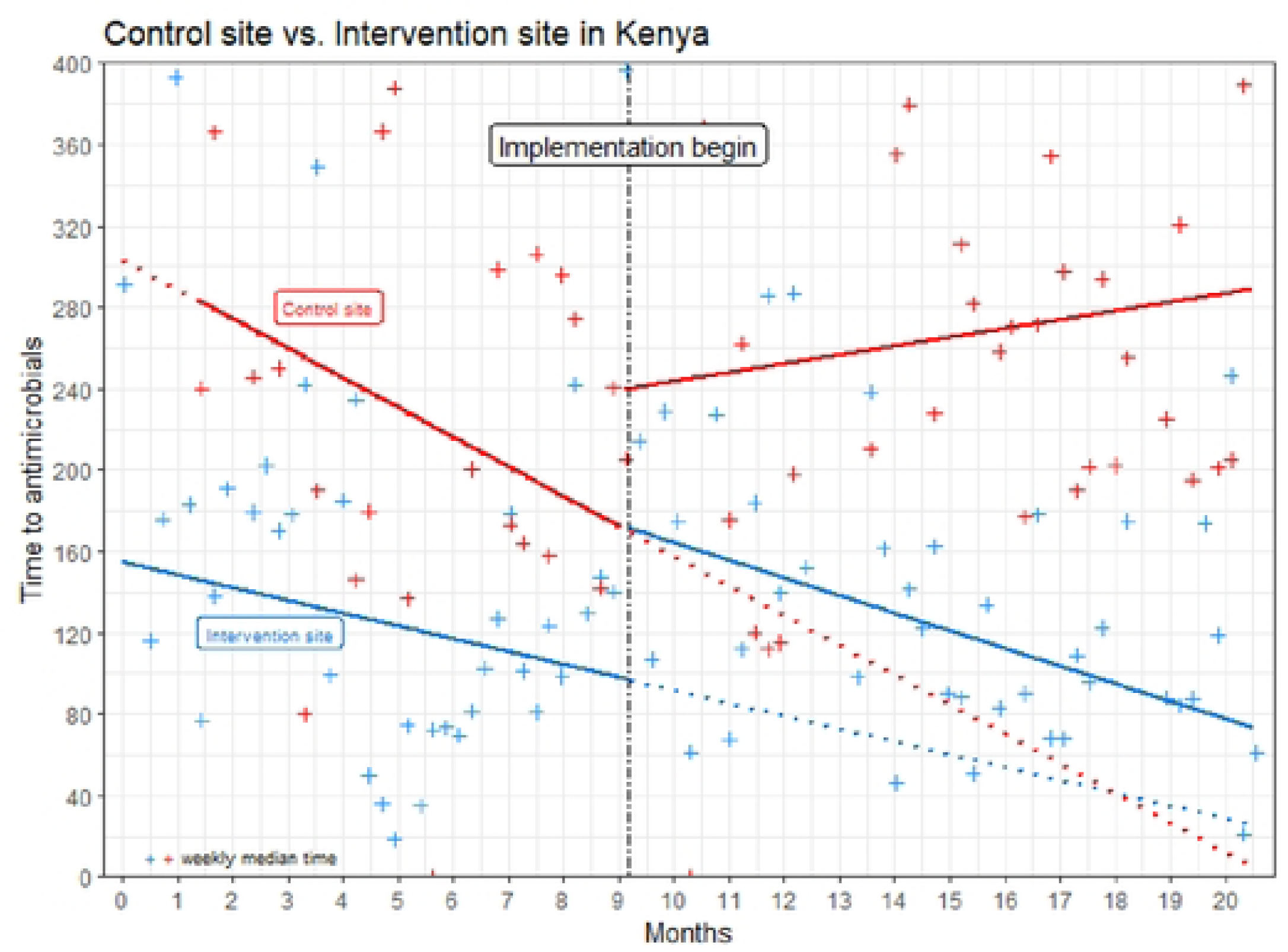
Interrupted time series analysis showing time to intravenous antimicrobials (IVA) in minutes at intervention and control sites in Kenya during baseline and implementation periods. The vertical dotted blackline indicates Smart Triage implementation. Secular trends are indicated by solid lines and counterfactual trends are indicated by dotted colored lines. Weekly median time to IVA is summarized by“+”.

**Table 1:** Segmented regression analysis of time to intravenous antimicrobial administration (IVA) at intervention and control sites during the implementation period.

|  | Kenya Estimates (95% CI) | Uganda Estimates (95% CI) |
| --- | --- | --- |
| Intervention site |  |  |
| Start of period | 171 (126 to 216) | 234 (200 to 267) |
| Slope (mins/week) | -2.0 (-0.3 to -3.7) | 0.3 (-1.4 to 2.0) |
| End of period | 73 (44 to 102) | 246 (213 to 280) |
| Control site |  |  |
| Start of period | 239 (138 to 341) | 222 (183 to 261) |
| Slope (mins/week) | 1.0 (-4.0 to 6.0) | -0.2 (-1.3 to 0.9) |
| End of period | 289 (214 to 364) | 209 (163 to 255) |
| Comparison between intervention site and control site |  |  |
| Difference in slopes (mins/week) | -3 (-8 to 2) | -0.1 (-0.2 to 1) |
| Difference at the beginning of period (mins) | -68 (-125 to -12) | 12 (6 to 17) |
| Difference at the end of period (mins) | -216 (-262 to -170) | 33 (25 to 50) |

Note: values are in minutes

In Uganda, the baseline time to IVA decreased by 3 minutes (2%, 95% CI: -12 to 5) at the intervention site (Figure 3). During the intervention, the time to IVA initially decreased by 19 min (8%, 95% CI: 19-21) compared to the end of baseline, but this was not sustained. The median time to IVA during the baseline period was 16 min (6%, 95% CI: 6 to 40) more than the implementation phase. Time to IVA at the Uganda control site decreased by 13 min (6%, 95% CI: 6 to 20) during the implementation phase, but there was no statistically significant difference between intervention and control sites (Table 2). We have no baseline data from the Uganda control site, so we could not evaluate the trend over time for the entire study period.

**Figure 3.**
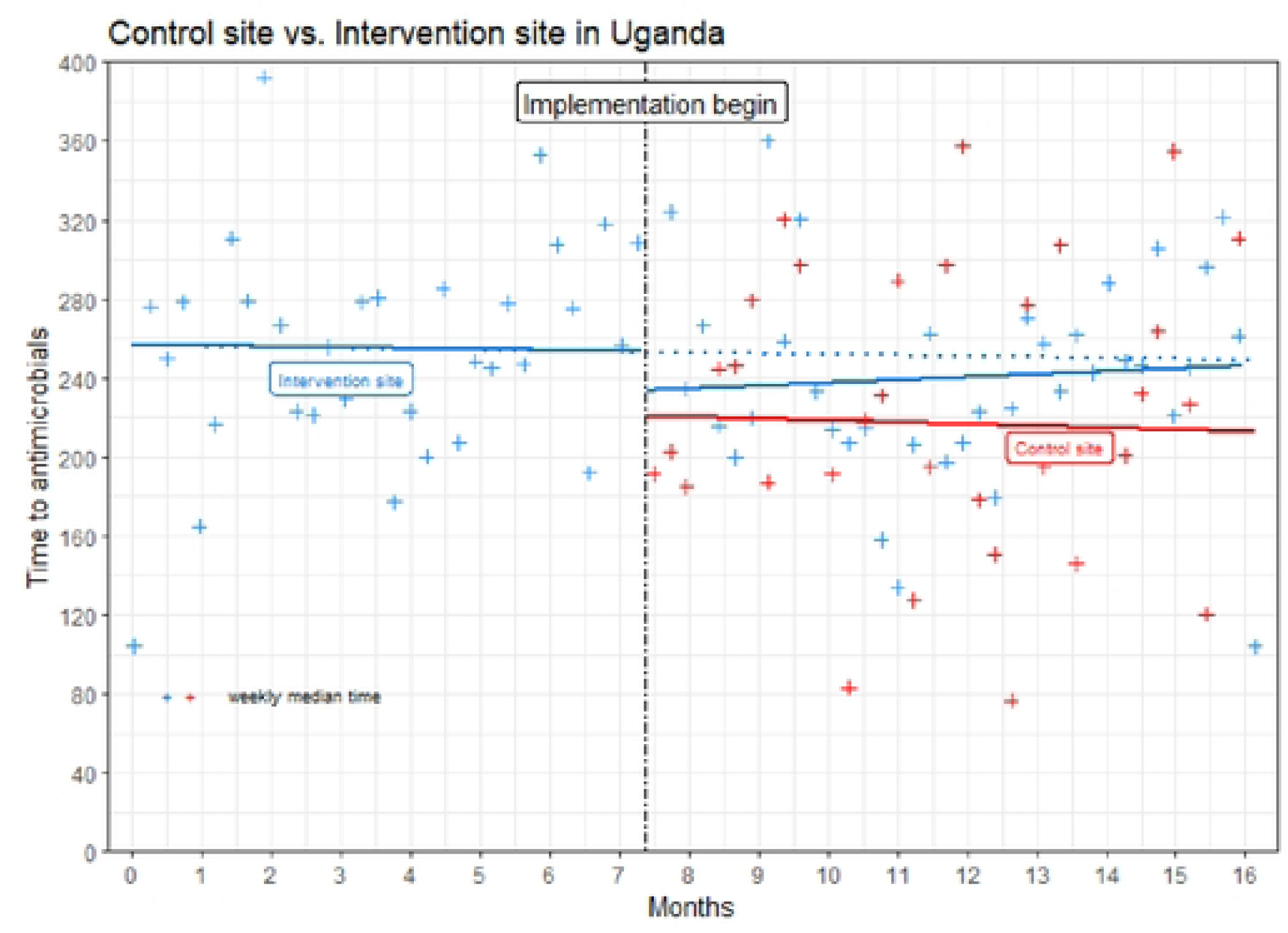
Interrupted time series analysis showing time to Intravenous antimicrobials (IVA) In minutes at Intervention and control sites in Uganda during baseline and implementation periods. The vertical dotted black line indicates Smart Triage implementation. Secular trends are indicated by solid lines and counterfactual trends are indicated by dotted colored lines. Weekly median time to IVA is summarized by “+”. Due to delays related to COVID-19, the Ugandan control site was initiated during the implementation phase at the intervention site.

**Table 2:** Uganda: enrolment, time to IVA, mortality, admissions and readmissions.

| Uganda | Control compared to intervention sites during implementation |  |  |  | Baseline compared to implementation at the intervention site |  |  |
| --- | --- | --- | --- | --- | --- | --- | --- |
|  | Intervention site | Control site | % diff | Effect size<sup>b</sup> | Baseline | % diff | Effect size<sup>c</sup> |
| Enrolled, N | 1903 | 2855 |  |  | 1408 |  |  |
| Time to IVA, median (IQR) | 239 (171-303) | 212 (129-295) | 13 | 26.78 (0.88, 52.69) | 255 (186-348) | -6 | -16.22 (-38.92, 6.48) |
| Admitted, n (%) | 288 (15.2) | 207 (7.3) | 108 | OR: 2.28 (1.88-2.75) | 326 (23.2) | -34 | OR: 0.59 (0.50-0.71) |
| IVA, n (%) | 268 (14.1) | 196 (6.9) | 104 | OR: 2.22 (1.83, 2.70) | 294 (20.9) | -33 | OR: 0.62 (0.52, 0.75) |
| IVA in admitted cases, n (%)<sup>a</sup> | 268 (100) | 194 (99.0) | 1 | OR: 0.90 (0.44-1.85) | 291 (99.0) | 1 | OR: 1.61 (0.91-2.86) |
| Time to IVA emergency, median (IQR) | 202 (155-277) | 195 (122-277) | 4 | 3.73 (-33.53, 40.99) | 244 (173-320) | -17 | -41.60 (-70.56, -12.64) |
| Time to IVA priority, median (IQR) | 266 (205-328) | 220 (120-299) | 21 | 43.05 (-43.32, 129.43) | 257 (194-377) | 4 | 8.98 (-26.88, 44.85) |
| Readmitted, n (%) | 27 (1.4) | 17 (0.6) | 57 | OR: 2.44 (1.33-4.49) | 37 (2.7) | -48 | OR: 0.53 (0.32-0.87) |
| Mortality within 7 days, n (%) | 4 (0.2) | 2 (0.07) | 186 | OR: 3.05 (0.56-16.67) | 11 (0.8) | -75 | OR: 0.27 (0.08-0.84) |

Note: <sup>a</sup>the denominator is total admitted; <sup>b</sup> reference is control site, OR is obtained by fitting univariate logistic regression with first column as the outcome and site as the independent variable; <sup>c</sup>reference is baseline, OR is obtained by fitting univariate logistic regression model with phase as the independent variable; IVA= intravenous antimicrobial administration; diff=difference; OR=odds ratio

In Kenya, there was a reduction in admission rates between the baseline and intervention of 47% (OR: 0.51, 95% CI: 0.42-0.62, p<0.0001) (Table 3). Admission rates in the control site were unchanged (4.7%). IVA use was also reduced at the intervention site compared to the baseline by 47% (OR: 0.51 CI: 0.41-0.63) (Table 3). IVA use in the Kenya control site was also unchanged (2.9%). Mortality within 7 days was also reduced by 25% (OR: 0.29, 95% CI: 0.14-0.59) compared to the baseline period.

**Table 3:** Kenya: Enrolment, time to IVA, mortality, admissions and readmissions at sites in Kenya.

| Kenya | Control compared to intervention sites at baseline |  |  |  | Control compared to intervention sites during implementation |  |  |  | Intervention site: Baseline compared to implementation at the intervention site |  |
| --- | --- | --- | --- | --- | --- | --- | --- | --- | --- | --- |
|  | Intervention site | Control site | % diff | Effect size<sup>b</sup> | Intervention site | Control | % diff | Effect size<sup>b</sup> | % diff | Effect size<sup>c</sup> |
| Enrolled, N | 2856 | 2520 |  |  | 3428 | 3177 |  |  |  |  |
| Time (min) to IVA, median (IQR) | 115 (68-203) | 239 (160-388) | -52 | -125 (-168, -81) | 123 (68-213) | 261 (193-392) | -53 | -139 (-179, -99) | 7 | 8 (-21, 37) |
| Admitted, n (%) | 255 (9.0) | 119 (4.7) | 90 | OR: 1.99 (1.59-2.49) | 164 (4.78) | 148 (4.7) | 3 | OR: 1.03 (0.82-1.29) | -47 | OR: 0.51 (0.42-0.62) |
| IVA, n (%) | 218 (7.6) | 73 (2.9) | 162 | OR: 2.77 (2.11, 3.63) | 138 (4.03) | 92 (2.9) | 39 | OR: 1.41 (1.08, 1.84) | -47 | OR: 0.51 (0.41, 0.63) |
| IVA in admitted cases, n (%)<sup>a</sup> | 193 (88.5) | 69 (94.5) | -6 | OR: 2.26 (1.42-3.58) | 108 (78.3) | 86 (93.5) | -16 | OR: 1.39 (0.88-2.21) | -12 | OR: 0.62 (0.40-0.95) |
| Time to IVA emergency cases, median (IQR) | 119 (65-185) | 196 (149-366) | -65 | -79 (-136, -21) | 101 (58-162) | 262 (201-381) | -61 | -161 (-209, -113) | -15 | -18 (-46, 10) |
| Time (min) to IVA priority cases, median (IQR) | 108 (59-235) | 303 (175-480) | -64 | -191 (-272, -109) | 169 (68-246) | 258 (179-354) | -34 | -87.63 (-181, 5) | 56 | 62 (-2, 127) |
| Readmitted, n (%) | 20 (0.7) | 13 (0.5) | 37 | OR: 2.39 (1.26-4.53) | 21 (0.6) | 27 (0.9) | 28 | OR: 0.72 (0.41-1.28) | -13 | OR: 0.88 (0.47-1.62) |
| Mortality within 7 days, n (%) | 29 (0.4) | 11 (1.0) | -60 | OR: 2.34 (1.17-4.67) | 10 (0.3) | 10 (0.3) | -6 | OR: 0.93 (0.39-2.24) | -25 | OR: 0.29 (0.14-0.59) |

Note: <sup>a</sup>denominator is total admitted; <sup>b</sup>reference is control site, OR is obtained by fitting univariate logistic regression with first column as the outcome and site as the independent variable;; <sup>c</sup>reference is baseline, OR is obtained by fitting univariate logistic regression model with phase as the independent variable; IVA= intravenous antimicrobial administration; diff=difference; OR=odds ratio

In Uganda, the reduction in admission rate between baseline and intervention was 34% (OR: 0.59, 95% CI 0.50-0.71) (Table 2). IVA use was reduced by 33% (OR: 0.62 CI: 0.52-0.75) (Table 2). Mortality within 7 days was reduced by 75% (OR: 0.27, 95% CI: 0.08-0.84) at the intervention sites compared to the baseline period.

In addition to the overall reduction in admission rates, there was a reduction in priority admission rates of 64% (OR: 0.35, 95% CI: 0.22-0.55) in Kenya (Table S4) and 32% (OR: 0.62, 95% CI: 0.47-0.83) in Uganda (Table S5). There was a 19% (OR: 0.79 95% CI: 0.54-1.17) reduction in the admission rates of priority cases at the Kenya control site compared to the baseline. Emergency cases who presented to the Kenya intervention site showed a 161 min (62%, 95% CI: 113 to 209) reduction in time to IVA when compared to the Kenya control site (Table S4). In Uganda, the reduction was 42 min (17%; 95% CI: 13 to 71) compared to the baseline period (Table S5).

## Discussion

We report the results of a controlled interrupted time series analysis implementing the Smart Triage platform to improve the time to treatment. We found an inconsistent reduction in the time to IVA. The time to IVA at the start of baseline was much lower in Kenya than in Uganda, with a much larger reduction in time to IVA during the baseline compared to the intervention and control sites in Uganda. In Kenya, we observed an immediate increase in time to IVA at both our intervention and control sites between the end of the baseline and the start of the implementation phase. This effect could be explained by the timing of the Smart Triage implementation, which started during the December holiday period when staff shortages was common and there was staff changeover. In contrast, the sustained improvement observed at the intervention site versus the control site supports implementation success at this site. In Uganda at the start of the intervention, the control site had a lower time to IVA than the intervention site and continued to improve over time while the intervention site worsened. The comparison between sites is compromised by delayed initiation of the control site due to the COVID-19 pandemic, the lack of simultaneous baseline data and the differences between two facilities in different countries with different confounders (26).

The consistent reduction of time to IVA administration at all sites during the baseline period would indicate a system change associated with baseline data collection. A similar contamination has been observed in cluster randomized trials of digital health interventions that mask the impact of the intervention (27, 28). The downward slope in time to IVA during the baseline period would also suggest that this effect had not reached steady state.

What was surprising in our study was the decrease in treatment and admission rates in both Uganda and Kenya. This finding may be due to adherence to triage criteria in almost all patients and less reliance on personal preferences among clinicians. However, without more robust long-term follow-up (>7 days) of all children after discharge we cannot be certain that the lower admission rates reflect better triage practices. However, we did not see an increase in antimicrobial utilization or readmission rates. A similar reduction in antimicrobial utilization has been shown in a randomized trial using an electronic algorithm-guided management platform (29). These findings warrant additional investigation into the impact on longer-term mortality following triage as post-discharge mortality is common and may also be more prevalent in patients who were not admitted (30).

The finding of a decrease in mortality by 25% (OR: 0.29, 95% CI:0.14-0.59) in Kenya and by 75% (OR: 0.27, 95% CI: 0.08-0.84) in Uganda compared to the baseline period is similar to an effect that has been reported in the US in a large 19 hospital American Academy of Paediatrics Paediatric Severe Sepsis Collaborative that were focused on sepsis QI in emergency department care (31). In these collaboratives, there were shared criteria for sepsis, standardized screening tools and electronic health record (EHR) embedded tools (such as triage screens and sepsis order sets). Improvements were noted in time to first clinical assessment, fluid bolus and antibiotics, with an associated improvement in 30-day all-cause mortality from 2.3% to 1.4%. Interestingly, even sites newer to improvement work demonstrated process metric improvement as well as mortality reductions, corroborating the benefits of an ‘all teach - all learn’ QI collaborative methodology (31, 32).

Our findings of inconsistent improvement are not surprising because the uptake and effectiveness of clinical decision support tools have had mixed success due to complexities in implementation contexts (15, 33). Beyond the setting, technology-supported innovations face additional complexity in implementation that stems from each aspect of the innovation. These complexities include addressing the clinical condition (especially when poorly characterized like sepsis), interacting technology components, and obtaining organizational, user and patient buy-in (34). These factors are amplified in poorly resourced settings which are plagued by overcrowding and understaffing in emergency departments, inadequate resources for timely laboratory testing and imaging, inadequate stock of antibiotics, and inadequate overall resources for paediatric sepsis management (35) (36) (16). Thus, to be effective, digital interventions addressing triage and QI must consider individual and systems-related factors and processes that influence facility performance (37–39).

Sepsis care in children is complex. Consideration must be given to biology of both pathogens (malaria, dengue, bacteria and viruses) and the patient including age and genetics; along with seasonality of infections, resource availability (personnel, drugs, diagnostic tools) and access, timing of presentation and progress of deterioration and co-morbidities such as malnutrition and HIV (40). Care occurs in facilities which are complex, dynamic environments continuously influenced by process inputs, organizational structure and resources, culture and individual motivations (41). In this milieu of constant shifts and changes it is not surprising that QI is difficult to initiate and sustain. An assumption of the interrupted time series methodology is that the baseline trend is not changing during the intervention (42). By selecting a process indicator and prioritizing randomization and prospective enrollment on arrival at the facility; we hoped to account for confounding related to variable case mix and severity, however this may not be the case.

The time to IVA was chosen because it is the most important factor in improving outcomes from sepsis and easy to track and compare between hospitals (43). However, IVA is vulnerable to changes in resource availability (44, 45). For example, using the data-driven QI process, we were able to identify that delivery of timely treatment was influenced not just by identification of emergency cases, but also staff availability, drug and reagent shortages, and lab equipment for diagnostics and caregivers’ ability to pay for the drug and other sundries. These issues were exacerbated by the COVID-19 pandemic and persisted throughout the study period. The best chance for success, therefore, relies on a holistic approach which not only addresses the clinical elements of the treatment guideline but also include supply chain management, maintenance policies and coevolution of health systems infrastructure with pediatric triage and sepsis guidelines (26, 46, 47).

## Strengths and Limitations

The major limitation of this study is that the methodology chosen was unable to account for the significant confounders during data collection such as the COVID-19 pandemic and significant disruptions in supplies and staffing. We were also limited by the lack of synchronous data during the baseline in Uganda. We underestimated the impact of data collection during the baseline on time to IVA and would have preferred to have a longer period of observation to allow for traction in QI initiatives and account for seasonality.

The strengths of the study are completeness of data, and the adjustment of the intervention using small iterative changes (responsive feedback), co-developed with the QI teams on site. We actively addressed issues related to usability, training and staff rotations through continued training, resources, and job aides. Sustainability and continued staff-led problem solving is supported through an ongoing QI program and partnerships with implementors.

## Conclusion

Smart Triage was successfully implemented for routine use in two resource-constrained settings. The improvement in time to IVA was inconsistent between sites and impacted by numerous health system factors. However, the reduction in admission and treatment times and the reduction in mortality are benefits worthy of further investigations. We have also highlighted the significant challenges in undertaking clinical evaluation of digital health tools in complex real-world clinical settings. The results can be leveraged to understand delivery gaps, strengthen implementation strategy and methodology and inform future adaptions of the Smart Triage platform.

## Data Availability

Study materials (protocol, data collection tools, data dictionary, software, analysis code, and metadata) are publicly available through the Pediatric Sepsis Data CoLaboratory’s (Sepsis CoLab) Dataverse on https://borealisdata.ca/dataverse/ST_implementation . Due to the sensitive nature of clinical data and the potential risk for re-identification of research participants, the de-identified dataset is available through moderated access on the Sepsis CoLab Dataverse on https://borealisdata.ca/dataverse/ST_implementation and through the KWTRP Research Data Repository Dataverse https://kemri-wellcome.org/dataverse/. Access to these data will be granted on a case-by-case basis following approval from the authors and the Data Governance Committees.

## Acknowledgements

We are grateful for the quality improvement efforts of facility staff and leadership at Jinja Regional Referral Hospital and Mbagathi County Hospital. Further, we would like to thank members of the Smart Triage research team based at Walimu, Uganda; Kenya Medical Research Institute, Kenya; and the Institute for Global Health, Canada. This includes but is not limited to, Savio Mwaka, Clare Komugisha, Bamwesigye Emmanuel, Annet Mary Nabweteme, Busense Bosco, Kisaame Meshack Moshin, Kantono Miria, Nakasagga Barbra, Monero Angel, Isaac Omara, Komakech Francis, Patricia Aloya, Joan Lamagi, Priscilla Antimango, Reagan Obalim, Angela Wanjiru, Anastasia Gathigia, Felix Kimani, Mercy Mutuku, Emmah Kinyanjui, Esther Muthoni, Faith Wairimu, Sidney Kipkorir, Verah Karasi, Victor Achiro, Kevin Bosek, Brian Ochieng, John Mboya, Deborah Lester, Jessica Rigg, Katija Pallot, Parnian Hosseini, Alishah Mawji and Edmond C K Li.

## References

1. Liu L, Oza S, Hogan D, Chu Y, Perin J, Zhu J, et al. Global, regional, and national causes of under-5 mortality in 2000–15: an updated systematic analysis with implications for the Sustainable Development Goals. The Lancet. 2016;388(10063):3027–35.

2. Kissoon N, Reinhart K, Daniels R, Machado MFR, Schachter RD, Finfer S. Sepsis in Children: Global Implications of the World Health Assembly Resolution on Sepsis. Pediatr Crit Care Med. 2017;18(12):e625–e7.

3. Barasa EW, Ayieko P, Cleary S, English M. A Multifaceted Intervention to Improve the Quality of Care of Children in District Hospitals in Kenya: A Cost-Effectiveness Analysis. PLOS Medicine. 2012;9(6):e1001238.

4. Mittal Y, Sankar J, Dhochak N, Gupta S, Lodha R, Kabra SK. Decreasing the Time to Administration of First Dose of Antibiotics in Children With Severe Sepsis. J Healthc Qual. 2019;41(1):32–8.

5. Koenig C, Schneider C, Morgan JE, Ammann RA, Sung L, Phillips B. Association of time to antibiotics and clinical outcomes in patients with fever and neutropenia during chemotherapy for cancer: a systematic review. Support Care Cancer. 2020;28(3):1369–83.

6. Weiss SL, Peters MJ, Alhazzani W, Agus MSD, Flori HR, Inwald DP, et al. Surviving Sepsis Campaign International Guidelines for the Management of Septic Shock and Sepsis-Associated Organ Dysfunction in Children. Pediatric Critical Care Medicine. 2020;21(2):e52–e106.

7. Moorthy GS, Pung JS, Subramanian N, Theiling BJ, Sterrett EC. Causal Association of Physician-in-Triage with Improved Pediatric Sepsis Care: A Single-Center, Emergency Department Experience. Pediatr Qual Saf. 2023;8(3):e651.

8. Buys H, Muloiwa R, Westwood C, Richardson D, Cheema B, Westwood A. An adapted triage tool (ETAT) at red cross war memorial children’s hospital medical emergency unit, Cape Town: an evaluation. South African Medical Journal. 2013;103(3):161–5.

9. World Health O. Emergency triage assessment and treatment (ETAT). Geneva: World Health Organization; 2005.

10. Rominski S, Bell SA, Oduro G, Ampong P, Oteng R, Donkor P. The implementation of the South African Triage Score (SATS) in an urban teaching hospital, Ghana. African Journal of Emergency Medicine. 2014;4(2):71–5.

11. Bryce J, Victora CG, Habicht J-P, Black RE, Scherpbier RW, on behalf of the MCEITA. Programmatic pathways to child survival: results of a multi-country evaluation of Integrated Management of Childhood Illness. Health Policy and Planning. 2005;20(suppl_1):i5–i17.

12. Hategeka C, Mwai L, Tuyisenge L. Implementing the Emergency Triage, Assessment and Treatment plus admission care (ETAT+) clinical practice guidelines to improve quality of hospital care in Rwandan district hospitals: healthcare workers’ perspectives on relevance and challenges. BMC health services research. 2017;17(1):1–12.

13. Hansoti B, Jenson A, Keefe D, De Ramirez SS, Anest T, Twomey M, et al. Reliability and validity of pediatric triage tools evaluated in Low resource settings: a systematic review. BMC Pediatr. 2017;17(1):37.

14. Schultz MJ, Dünser MW, Dondorp AM, Adhikari NK, Iyer S, Kwizera A, et al. Current challenges in the management of sepsis in ICUs in resource-poor settings and suggestions for the future. Sepsis management in resource-limited settings. 2019:1–24.

15. Beynon F, Guérin F, Lampariello R, Schmitz T, Tan R, Ratanaprayul N, et al. Digitalizing Clinical Guidelines: Experiences in the Development of Clinical Decision Support Algorithms for Management of Childhood Illness in Resource-Constrained Settings. Global Health: Science and Practice. 2023.

16. Melendez E, Bachur R. Quality improvement in pediatric sepsis. Current Opinion in Pediatrics. 2015;27(3):298–302.

17. Clark M, Spry E, Daoh K, Baion D, Skordis-Worrall J. Reductions in inpatient mortality following interventions to improve emergency hospital care in Freetown, Sierra Leone. PLoS One. 2012;7(9):e41458.

18. Mawji A, Li E, Chandna A, Kortz T, Akech S, Wiens MO, et al. Common data elements for predictors of pediatric sepsis: A framework to standardize data collection. PLoS One. 2021;16(6):e0253051.

19. Mawji A, Li E, Dunsmuir D, Komugisha C, Novakowski SK, Wiens MO, et al. Smart triage: Development of a rapid pediatric triage algorithm for use in low-and-middle income countries. Front Pediatr. 2022;10:976870.

20. Mawji A, Li E, Komugisha C, Akech S, Dunsmuir D, Wiens MO, et al. Smart triage: triage and management of sepsis in children using the point-of-care Pediatric Rapid Sepsis Trigger (PRST) tool. BMC Health Services Research. 2020;20(1):493.

21. Novakowski SK, Kabajaasi O, Kinshella MW, Pillay Y, Johnson T, Dunsmuir D, et al. Health worker perspectives of Smart Triage, a digital triaging platform for quality improvement at a referral hospital in Uganda: a qualitative analysis. BMC Pediatr. 2022;22(1):593.

22. Li ECK, Tagoola A, Komugisha C, Nabweteme AM, Pillay Y, Ansermino JM, et al. Cost-effectiveness analysis of Smart Triage, a data-driven pediatric sepsis triage platform in Eastern Uganda. BMC Health Serv Res. 2023;23(1):932.

23. Karlen W, Gan H, Chiu M, Dunsmuir D, Zhou G, Dumont GA, et al. Improving the Accuracy and Efficiency of Respiratory Rate Measurements in Children Using Mobile Devices. PLOS ONE. 2014;9(6):e99266.

24. Gan H, Karlen W, Dunsmuir D, Zhou G, Chiu M, Dumont GA, et al. The Performance of a Mobile Phone Respiratory Rate Counter Compared to the WHO ARI Timer. J Healthc Eng. 2015;6(4):691–703.

25. Harris PA, Taylor R, Thielke R, Payne J, Gonzalez N, Conde JG. Research electronic data capture (REDCap)—a metadata-driven methodology and workflow process for providing translational research informatics support. Journal of biomedical informatics. 2009;42(2):377–81.

26. English M. Improving emergency and admission care in low-resource, high mortality hospital settings—not as easy as A, B and C. Health Policy and Planning. 2021;37(6):808–10.

27. Payne B, Kinshella MLW, Bawani S, Sheikh S, Hoodbhoy Z, Charanthimath U, et al. 216. Evaluation of the PIERS on the MOVE mobile health tool for pre-eclampsia triage: The users’ perspective. Pregnancy Hypertension. 2018;13:S101.

28. Payne B, Dunsmuir D, Qureshi R, Sevene E, Munguambe K, Goudar S, et al. 218. Implementation of the PIERS on the Move mobile health app in the Community Level Interventions for Pre-eclampsia trials. Pregnancy Hypertension. 2018;13:S101–S2.

29. Tan R, Kavishe G, Luwanda LB, Kulinkina AV, Renggli S, Mangu C, et al. A digital health algorithm to guide antibiotic prescription in pediatric outpatient care: a cluster randomized controlled trial. Nature Medicine. 2024;30(1):76–84.

30. Chaudhry M, Knappett M, Nguyen V, Trawin J, Mugisha NK, Kabakyenga J, et al. Pediatric post-discharge mortality in resource-poor countries: A protocol for an updated systematic review and meta-analysis. Plos one. 2023;18(2):e0281732.

31. Depinet H, Macias CG, Balamuth F, Lane RD, Luria J, Melendez E, et al. Pediatric Septic Shock Collaborative Improves Emergency Department Sepsis Care in Children. Pediatrics. 2022;149(3).

32. Britto MT, Fuller SC, Kaplan HC, Kotagal U, Lannon C, Margolis PA, et al. Using a network organisational architecture to support the development of Learning Healthcare Systems. BMJ Qual Saf. 2018;27(11):937–46.

33. Keitel K, D’Acremont V. Electronic clinical decision algorithms for the integrated primary care management of febrile children in low-resource settings: review of existing tools. Clin Microbiol Infect. 2018;24(8):845–55.

34. Greenhalgh T, Wherton J, Papoutsi C, Lynch J, Hughes G, A’Court C, et al. Analysing the role of complexity in explaining the fortunes of technology programmes: empirical application of the NASSS framework. BMC Medicine. 2018;16(1):66.

35. Cruz AT, Lane RD, Balamuth F, Aronson PL, Ashby DW, Neuman MI, et al. Updates on pediatric sepsis. Journal of the American College of Emergency Physicians Open. 2020;1(5):981–93.

36. Souza DC, Jaramillo-Bustamante JC, Céspedes-Lesczinsky M, Quintero EMC, Jimenez HJ, Jabornisky R, et al. Challenges and health-care priorities for reducing the burden of paediatric sepsis in Latin America: a call to action. The Lancet Child & Adolescent Health. 2022;6(2):129–36.

37. Odetola FO, Freed G, Shevrin C, Madden B, McCormick J, Dombkowski K. In-hospital quality-of-care measures for pediatric sepsis syndrome. Pediatrics. 2017;140(2).

38. Schlapbach LJ, Kissoon N. Defining pediatric sepsis. JAMA pediatrics. 2018;172(4):313–4.

39. Dalwai M, Tayler-Smith K, Twomey M, Wallis L. Developing a reference standard for assessing paediatric triage scales in resource poor settings. African journal of emergency medicine. 2015;5(4):181–4.

40. Carrol ED, Ranjit S, Menon K, Bennett TD, Sanchez-Pinto LN, Zimmerman JJ, et al. Operationalizing Appropriate Sepsis Definitions in Children Worldwide: Considerations for the Pediatric Sepsis Definition Taskforce. Pediatr Crit Care Med. 2023;24(6):e263–e71.

41. English M, Irimu G, Wamae A, Were F, Wasunna A, Fegan G, et al. Health systems research in a low-income country: easier said than done. Archives of Disease in Childhood. 2008;93(6):540–4.

42. Hategeka C, Ruton H, Karamouzian M, Lynd LD, Law MR. Use of interrupted time series methods in the evaluation of health system quality improvement interventions: a methodological systematic review. BMJ Global Health. 2020;5(10):e003567.

43. Evans IV, Phillips GS, Alpern ER, Angus DC, Friedrich ME, Kissoon N, et al. Association between the New York sepsis care mandate and in-hospital mortality for pediatric sepsis. Jama. 2018;320(4):358–67.

44. English M, Ntoburi S, Wagai J, Mbindyo P, Opiyo N, Ayieko P, et al. An intervention to improve paediatric and newborn care in Kenyan district hospitals: Understanding the context. Implementation Science. 2009;4(1):42.

45. Moore GF, Audrey S, Barker M, Bond L, Bonell C, Hardeman W, et al. Process evaluation of complex interventions: Medical Research Council guidance. Bmj. 2015;350:h1258.

46. King C, Dube A, Zadutsa B, Banda L, Langton J, Desmond N, et al. Paediatric Emergency Triage, Assessment and Treatment (ETAT) - preparedness for implementation at primary care facilities in Malawi. Glob Health Action. 2021;14(1):1989807.

47. Bryce J, Victora CG, Habicht J-P, Vaughan JP, Black RE. The Multi-Country Evaluation of the Integrated Management of Childhood Illness Strategy: Lessons for the Evaluation of Public Health Interventions. American Journal of Public Health. 2004;94(3):406–15.

